# Dissecting the causal relationships between childhood-onset asthma and major mental disorders: a univariable and multivariable Mendelian randomization study

**DOI:** 10.1101/2023.10.09.23296789

**Authors:** Banghong Chen, Mingzhu Xue, Lan Zhang, Peng Ren

**Affiliations:** Data Science R&D Center of Yanchang Technology, Chengdu 610041, Sichuan, China; School of Life Sciences and Engineering, Southwest University of Science and Technology, Mianyang 621010, Sichuan, China

## Abstract

**Background:** Asthma with a childhood-onset is found to be associated with increased risk of severe mental illnesses in later life. However, the causal relationships between childhood-onset asthma and major mental disorders remained unclear.

**Methods:** We conducted a two-sample Mendelian randomization (MR) analysis to investigate the causal effects of childhood-onset asthma (n=327,670) on six major mental illnesses, including major depressive disorders (n=143,265), bipolar disorder (n=353,899), schizophrenia (n=130,644), anxiety (n=10,240), autism (n=46,350), and ADHD (n=225,534) using summary statistics of genome-wide association studies (GWAS). The inverse variance weighted (IVW) method, along with weighted median and MR-Egger were employed for the causal estimates. Multiple sensitivity analyses were conducted to examine the robustness of the estimates. Moreover, the direct effects of childhood-onset asthma on mental disorders after accounting for the effects of adult-onset asthma were evaluated through the multivariable MR (MVMR) analysis.

**Results:** We found that genetically determined childhood-onset asthma significantly increased the risk of depression (IVW OR=1.059, 95%CI:1.025-1.095, p=5.72e-04) and bipolar disorder (IVW OR=1,065, 95%CI:1.027-1.105, p=6.75e-04), but not associated with other mental disorders. Further MVMR analysis indicated that the causal relationships remained significant with the adjustment of adult-onset asthma. Interestingly, we found that childhood-and adult-onset asthma exerted distinct causal effects on depression and bipolar disorders. No significant heterogeneity and horizontal pleiotropy were found to influence the causal estimates.

**Conclusions:** MR analysis indicated a significant causal relationship between genetically determined childhood-onset asthma and increased risk of depression and bipolar disorder in later life. The causal effects of childhood-onset asthma were distinct to the adult-onset asthma. Further studies were warranted to investigate the mechanisms underlying the causal relationships.

## Introduction

Asthma is the most prevalent chronic respiratory disorder characterized by presence of airflow obstruction and clinical symptoms including wheeze, cough, and shortness of breath [1]. More than 14% of children worldwide have a diagnosis of asthma, which imposes substantial medical and psychosocial morbidity [2]. In particular, accumulating studies have indicated that asthma with a childhood-onset is associated with increased risk of severe mental illnesses in later life. Meta-analyses of observational studies have reported children with asthma had higher risk of developing subsequent major depressive disorder (MDD) [3, 4], bipolar disorder [4, 5], schizophrenia [5], anxiety [6], autism [7]and attention deficit hyperreactivity disorder (ADHD) [8, 9]. While various covariates have been controlled in these studies, the remaining uncontrolled confounding factors and reverse causation bias still limit the ability of elucidating the causal relationships from observational studies [10]. Understanding the causal relationships would provide insights into the disease pathogenesis, management, and therapeutics development.

Mendelian randomization (MR) is a promising approach that uses genetic proxies for potential risk factors to investigate the causal effects on the outcome diseases [11]. The MR method is less likely to be biased by unmeasured confounding factors and reverse causation, as the genetic variants were randomly passed from the parents to the offspring and fixed at conception. Therefore, the MR approach is conceptually analogous to a randomized controlled trial (RCT) study, while being more widely used and cost-effective [12]. A recent MR study has reported that asthma overall (without considering the age of disease onset) is not playing a causal role in the development of mental health diseases [13]. However, compelling studies have shown that asthma is a highly heterogeneous disease, in which childhood-onset asthma and adult-onset asthma differ with respect to etiology [14], severity [15], and underlying genetic risk factors [16, 17]. Therefore, whether childhood-onset asthma exerts causal effects on developing subsequent mental disorders remains unclear.

In this study, we conducted a two-sample MR analysis to investigate whether childhood-onset asthma could be causally associated with six major mental illnesses including depression, bipolar disorder, schizophrenia, anxiety, autism, and ADHD. Childhood-onset asthma (binary phenotype, onset before 12 years) and age of onset (contiguous phenotype) were used as the exposure variables respectively. Multiple sensitivity analyses were performed to ensure the robustness of the results. In addition, we performed multivariable MR (MVMR) analysis to estimate the independent effect of childhood-onset asthma on the significant mental disorders while adjusting for the effect of adult-onset asthma.

## Data and Methods

### Study design

Two-sample MR design was unitized to explore the causal relationship between childhood-onset asthma and risk of mental disorders. For the causal estimates to be valid, three MR assumptions need to be hold: 1. the genetic instruments are significantly associated with the exposure variables; 2. the genetic variants are not associated with confounding factors; 3. the genetic variants affect the outcome only through the exposure while not through any alternative pathways [18]. The overview design of the study and the fundamental MR assumptions were summarized in **Figure 1**.

**Figure 1.**
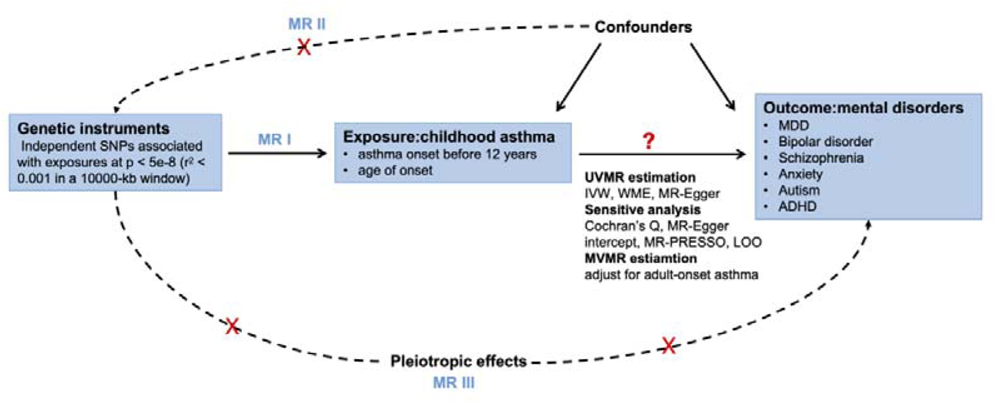
Overview of the study design and three MR assumptions. MR1: the genetic instruments are significantly associated with the exposure variables; MR2: the genetic variants are not associated with confounding factors; MR3: the genetic variants affect the outcome only through the exposure while not through any alternative pathways. MDD: Major Depressive Disorder; ADHD: attention deficit hyperreactivity disorder;

### Summary GWAS data

We extracted the summary association data from the latest GWAS study on childhood-onset and adult-onset asthma [16]. The study consists of 37,846 asthma patients and 318,237 controls from the UK Biobank data. Asthma patients with onset before 12 years were identified as childhood-onset cases (n=9,433) and those with onset between 26 and 65 years old as adult-onset cases (n=21,564). Individuals older than 38 years without an asthma diagnosis were included as controls (n=318,237). In the same study, we also extracted the top GWAS loci associated with the age-of-onset of asthma patients (n=37,846).

Similarly, we used summary statistics from the largest-to-date GWAS for mental disorders from the Psychiatric Genomics Consortium (PGC). Data generated from participants of European ancestry for MDD (45,591 cases and 97,674 controls) [19], bipolar disorder (40,463 cases and 313,436 controls) [20], schizophrenia (53,386 cases and 77,258 controls)[21], anxiety (2248 cases and 7992 controls)[22], autism (18381 cases and 27969 controls)[23], and ADHD (38,691 cases and 186,843 controls)[24]were extracted. To avoid the inflated type I error from the potentially sample overlap, we only kept the data without UKBB samples.

As all the summary-level GWAS data used for MR analyses were publicly available, no specific ethical approval is required. The information on the phenotype, sample size, and PubMed ID of the original study were summarized in **Supplementary Table 1.**

### Genetic instruments selection

To estimate the childhood-onset specific causal effects and minimize potential horizontal pleiotropy bias, we selected the single nucleotide polymorphisms (SNPs) that are specifically associated with childhood-onset asthma at the genome-wide significance threshold (p<5e-8), while not associated with adult-onset asthma (p>0.05). Then the independent SNPs were identified by clumping (r^2 < 0.001 in a 10000-kb window) the data based on European reference data from the 1000 Genomes Project [25]. Then the summary-level associations of selected instruments with mental disorders were extracted from the GWAS studies previously mentioned. The results were harmonized between exposure and outcome data according to the same effect allele. For palindromic SNPs, the reported allele frequency was checked to avoid potential strand flipping issues. Palindromic SNPs with minor allele frequency larger than 0.42 were removed due to ambiguity in inferring strand. The strength of the instrument (F statistics) was calculated as 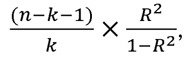 where n represents sample size, k represents the number of genetic variants, and R^2^ is the proportion of phenotypic variance explained by the genetic variants. All the instruments used in the study have F statistics larger than the empirical threshold of 10, indicating minimum week instrument bias [26]. The information on the genetic instruments used in this study can be found in **Supplementary Table 2.**

### Univariable MR analysis

Causal effects of childhood-onset asthma on mental disorders were estimated using the multiplicative random-effects inverse-variance weighted (IVW) method to account for potential heterogeneity [26]. Since IVW method is prone to be affected by the horizontal pleiotropy, we also analyzed the causal effects using two alternative MR methods including weighted median estimator and MR-Egger. Weighted median method will provide an unbiased estimate as long as at least 50% of instruments were valid [27], while MR-Egger will provide an unbiased MR estimate assuming the independence between the instrument and potential pleiotropy [28].

### Sensitivity analysis

Multiple sensitivity analyses were performed to examine the robustness of the results. Heterogeneity among SNPs was evaluated using the Cochran’s Q statistics. Leave-one-out analysis was used to examine whether the MR estimation was driven by specific genetic variant. The horizontal pleiotropy was assessed by the significance of MR-Egger intercept [28] and MR-PRESSO global test [29]. If a potential horizontal pleiotropy was detected, MR-PRESSO outlier test was performed to identify outliers. We then removed the outliers and assessed the causal relationships with the remaining SNPs. The causal relationship was considered significant if the following three conditions were met: a. the results of the IVW method passed the Bonferroni-corrected threshold (p<0.05/6=0.008); b. the directions of the estimates by the three methods (IVW, weighted median, and MR-Egger) were consistent; c. no significant horizontal pleiotropy was detected by MR-Egger intercept test (p>0.05) and MR-PRESSO global test (p>0.05).

### Multivariable MR analysis

Given that childhood-onset asthma and adult-onset asthma are correlated and have shared genetic risk factors [16, 17], we then performed a multivariable MR analysis (MVMR) to assess the direct effect of childhood-onset asthma on the mental disorders with adjustment of the effect of adult-onset asthma. We performed the MVMR analysis for the mental disorders that show significant causal relationships with childhood-onset asthma in the univariable MR analysis. The multivariable extension of the IVW method was used to estimate the direct effects of childhood-and adult-onset asthma on mental disorders. All the MR analyses were analyzed using “TwoSampleMR” package (version 0.5.6) [30]in R (version 4.0.0).

## Results

### Genetic instruments selection

Two exposures were included for the MR analysis: childhood-onset asthma (onset before 12 years) and age of onset. After the rigorous instrumental variables selection procedures, the number of valid genetic variants associated with exposures ranged from 9 to 19 depending on the different outcomes. The F-statistics of all the instruments were larger than 10 (ranged from 41.6 to 83.7), indicating the results were unlikely to be biased by the weak instrument **(Supplementary Table 2).**

### The causal effect of childhood-onset asthma on mental disorders by univariable MR analysis

Given the Bonferroni corrected threshold (p<0.05/6=0.008), the genetic proxy for childhood asthma was found to be significantly associated with an increased risk of depression (IVW OR=1.059, 95%CI:1.025-1.095, p=5.72e-04) and bipolar disorder (IVW OR=1,065, 95%CI:1.027-1.105, p=6.75e-04). The MR estimates were comparable when using alternative MR approaches, including weighted median and MR-Egger regression (**Figure 2**, **Table 1**). Consistently, we also found that each year earlier in genetically-predicted age of onset of asthma significantly increased depression risk (IVW OR=0.993, 95%CI:0.988-0.998, p=3.27e-03). There was also a negative trend between age of onset of asthma and bipolar disorder risk, however, the MR estimate was not significant. There was no evidence for significant causal effects of childhood asthma on other mental disorders including schizophrenia, anxiety, autism, and ADHD (all IVW p > 0.05) (**Supplementary Table 3).**

**Figure 2.**
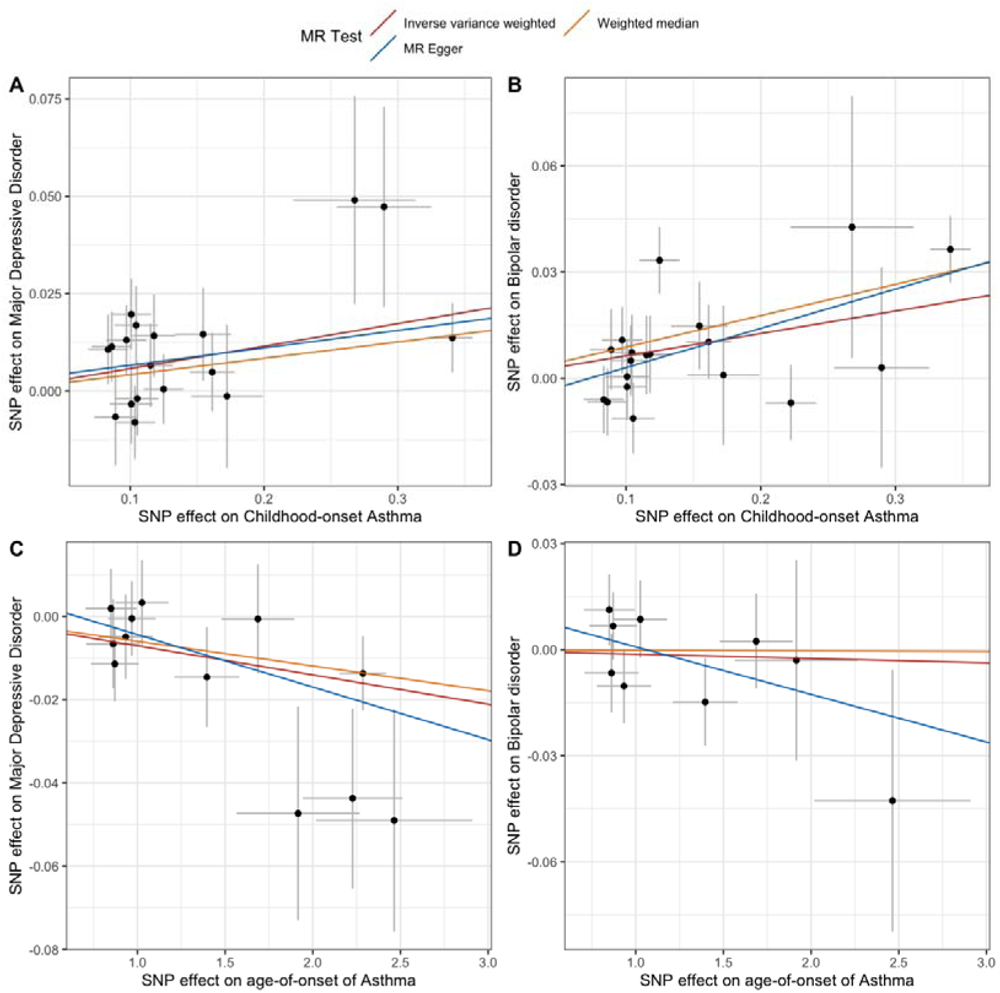
Scatter plot of the selected causal estimates. A. Scatter plot of the causal estimates between childhood-onset asthma and major depressive disorder; B. Scatter plot of the causal estimates between childhood-onset asthma and bipolar disorder; C. Scatter plot of the causal estimates between age-of-onset of asthma and major depressive disorder; D. Scatter plot of the causal estimates between age-of-onset of asthma and major depressive disorder.

**Table 1.**
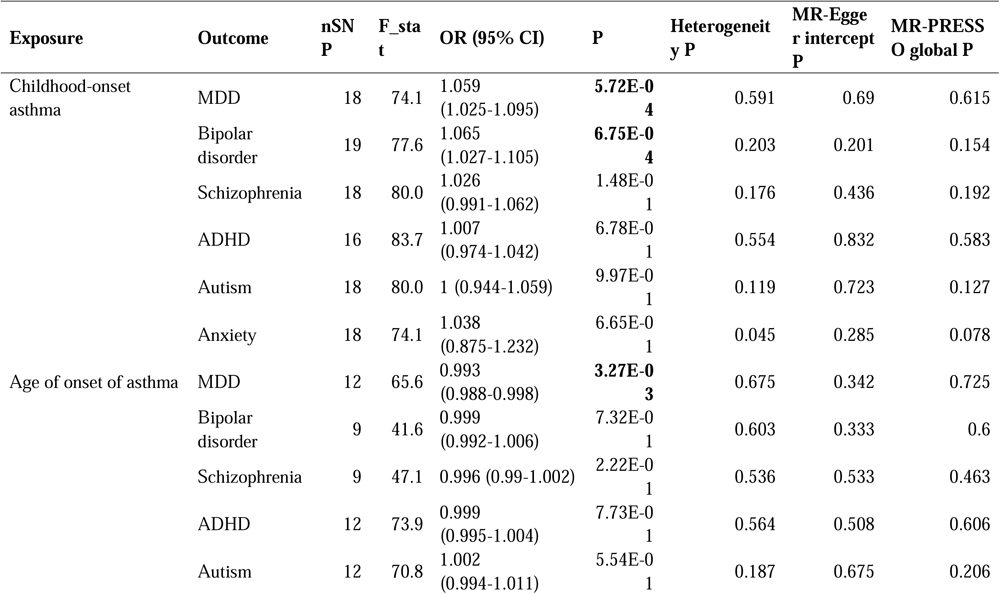
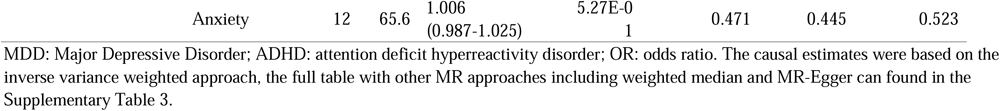
The causal effect of childhood-onset asthma on mental disorders by univariable MR analysis.

Cochran’s Q statistics indicated that no significant heterogeneity was detected in most of the MR estimates (p > 0.05). As for the existence of horizontal pleiotropy, both MR-Egger intercept test and MR-PRESSO test suggested that the results were less likely to be affected by the pleiotropic effects (all p values > 0.05) (**Table 1**). Furthermore, leave-one-out analysis supported that the MR estimates were not driven by a specific SNP (Supplementary Figure 1). Taken together, the results of sensitivity analyses ensured the robustness of the causal estimates.

### Distinct causal effects of childhood-and adult-onset asthma on mental disorders by multivariable MR analysis

As the childhood-and adult-onset asthma were correlated and shared genetic risk factors [16, 17], multivariable MR (MVMR) analysis was further performed to assess the direct effect of childhood-onset asthma on the significant mental disorders (i.e. depression and bipolar disorder) with adjustment of the effect of adult-onset asthma (**Figure 3**). Consistent with the results of univariable MR analysis, childhood-onset asthma was significantly associated with increased risk of depression (IVW OR=1.052, 95%CI: 1.020-1.084, p=1.25e-03) after accounting for adult-onset asthma. Interestingly, adult-onset asthma was found to exert a protective effect on depression (IVW OR=0.886, 95%CI: 0.817-0.962, p=3.68e-03) with adjustment of childhood asthma. Similarly, childhood-onset asthma was found to be significantly associated with increased risk of bipolar disorder (IVW OR=1.072, 95%CI: 1.021-1.127, p=5.66e-03), while there was a negative association between adult-onset asthma and risk of bipolar disorder (IVW OR=0.882, 95%CI: 0.772-1.006, p=0.062). Taken together, MVMR analysis indicated that childhood-and adult-onset asthma had distinct causal effects on depression and bipolar disorder.

**Figure 3.**
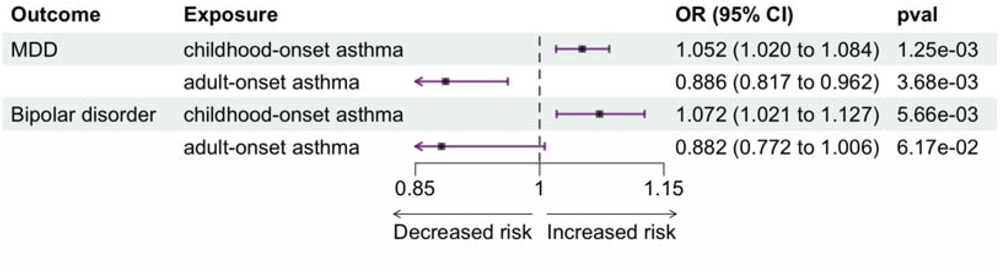
Multivariable MR analysis of the causal effects of childhood-and adult-onset asthma on major depressive disorder (MDD) and bipolar disorder.

## Discussions

We investigated the causal relationship between childhood asthma and six major mental disorders using two-sample MR approaches based on large-scale publicly available GWAS summary data. We found significant causal effects of childhood-onset asthma on increased risk of developing depression and bipolar disorder. Consistently, we also found that earlier age of onset of asthma was causally associated with increased of risk of depression. The causal relationship remained significant after accounting for the effect of adult-onset asthma. Interestingly, the adult-onset asthma was shown to exert opposite causal effect on mental illnesses (depression and bipolar disorder) compared to the childhood-onset asthma. To our best knowledge, this was the first MR study to dissect the distinct causal effects of childhood-and adult-onset asthma on mental health. Our findings would have important implications in understanding the disease pathogenesis and subtypes, improving the disease management, and developing effective therapeutics.

The link between between asthma and mental disorders had been well documented in observational studies [3–8], but the causality remained unclear. Previous studies using GWAS data suggested shared genetic liability between asthma overall (without considering the age of disease onset) and depression [31], but MR analysis didn’t show significant causal relationship [13, 32]. Given that childhood-and adult-onset asthma were increasingly being recognized as two different asthma subtypes, it is critical to separate the causal effects by the age of onset of disease. Our results indicated that genetically predicted childhood-onset asthma significantly increased the risk of depression and bipolar disorder, while adult-onset asthma was correlated with a lower risk of mental illness. Recent studies suggested that the disturbance of immune system by allergic disease like asthma might contribute to the development of psychiatric disorders [33, 34]. Previous MR studies also indicated that genetically predicted inflammatory markers, e.g. IL-6, C-reactive protein (CRP), were positively associated with risk of depression [35]and bipolar disorder [35]. The distinct causal effects of childhood-and adult-onset asthma indicated that the immune profiles of the two subtypes of asthma might be different. This is in line with the findings that adult-onset asthma tended to have a significant higher proportion of non-Th2 high patients than the children asthma patients [36], suggesting different biological pathways might be involved behind the two subtypes. Further studies on the molecular mechanisms through which childhood-and adult-onset asthma affect depression and bipolar disorder development were warranted.

Dissecting the causal relationship between asthma and mental disorders have multiple clinical implications. On the one hand, our results suggested the potential benefits of screening and/or early intervention among children asthma patients in reducing the risk of developing subsequent depression and bipolar disorders. On the other hand, the causal relationships would support the feasibility of repurposing existing asthma treatments for depression and bipolar disorder. A meta-analysis of randomized clinical trials indicated that the use of nonsteroidal anti-inflammatory drugs (NSAIDs) had beneficial effects in decreasing depression symptoms [37]. The effects of NSAIDs on bipolar disorder therapy was also evaluated [38]. More clinical trials were warranted to investigate the efficacy of anti-inflammation drugs in the treatments of mental disorders.

It is noted that there are several limitations to this study. First, the MR estimates were restricted to the European population due to the availability of the data, so the findings should not be extrapolated to the other ethnic groups. The causal estimates in general population warrants further studies. Second, the diagnoses of asthma in UK Biobank were predominantly self-reported, thus it is likely that the information of diagnosis and age-of-onset were misreported. However, the GWAS study was able to replicate most of the previously reported asthma liability loci [38], suggesting the results were robust to the potential data inaccuracies. Further studies should consider using large sample size cohort with clinically verified phenotypes to validate the causal estimations.

In conclusion, Mendelian randomization analysis indicated a significant causal relationship between genetically determined childhood-onset asthma and increased risk of depression and bipolar disorder in later life. The causal effects of childhood-onset asthma were distinct to the adult-onset asthma. Our findings would have important implications in better understanding the disease pathogenesis and subtypes, improving the disease management, and developing effective therapeutics.

## Declarations

### Ethics approval and consent to participate

Not applicable

## Consent for publication

Not applicable

## Availability of data and materials

The data used to support the findings of this study are included within the article.

## Conflict of interest

The authors declare that there are no conflicts of interest.

## Funding

The study did not receive any specific funding from funding agencies in the public, commercial or non-profit sectors.

## Author Contributions

All authors contributed to the study conception and design. **Banghong Chen**: Conceptualization, Methodology, Software, Writing-Original draft, Data curation, Visualization were performed; **Mingzhu Xue and Lan Zhang**: Investigation, Writing - Original Draft, Writing - Reviewing and Editing were performed; **Peng Ren**: Conceptualization, Supervision, Project administration were performed. All authors read and approved the final manuscript.

## Data Availability

The data used to support the findings of this study are included within the article.

## Acknowledgements

Not applicable

## Notes

### Competing Interest Statement

The authors have declared no competing interest.

