## Supplementary_materials for "Dissecting the causal relationships between childhood-onset asthma and major mental disorders: a univariable and multivariable Mendelian randomization study"

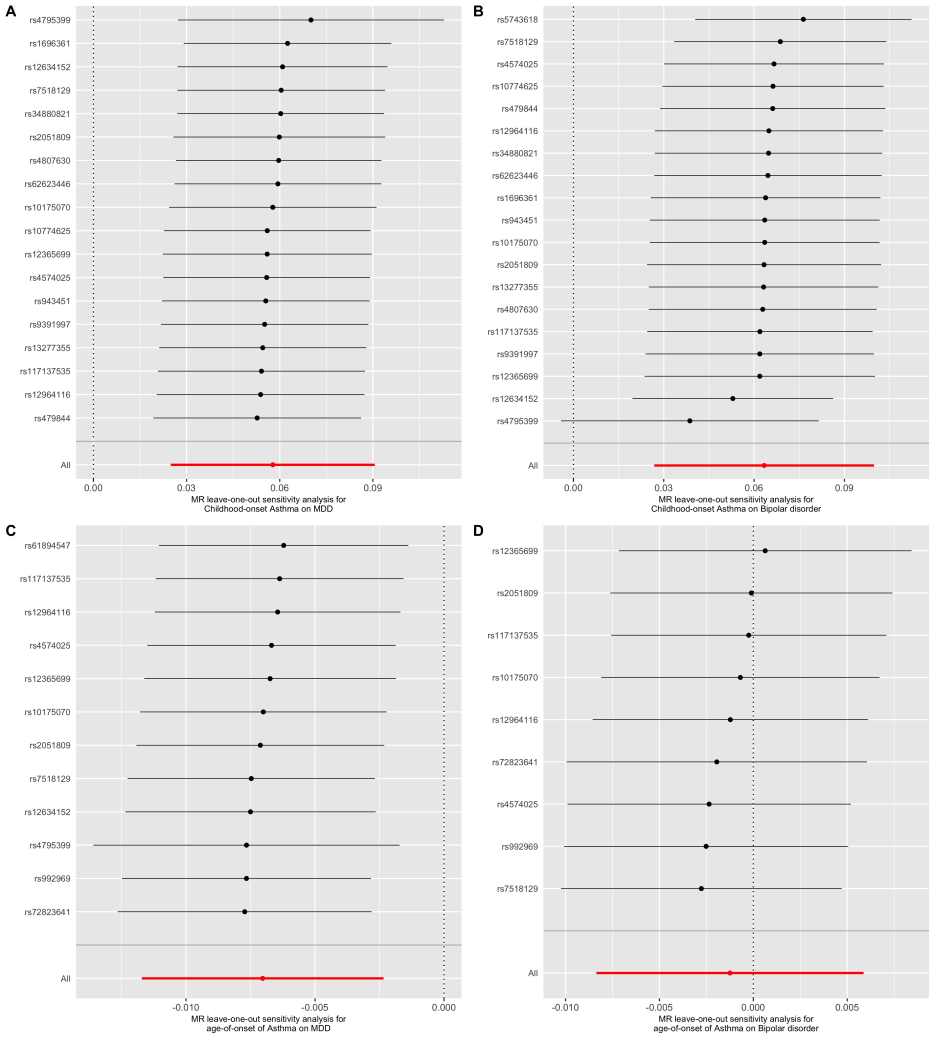

**Supplementary Figure 1. Leave-one-out (LOO) analysis**

A. LOO plot of the causal estimates between childhood-onset asthma and major depressive disorder; B. LOO plot of the causal estimates between childhood-onset asthma and bipolar disorder; C. LOO plot of the causal estimates between age-of-onset of asthma and major depressive disorder; D. LOO plot of the causal estimates between age-of-onset of asthma and major depressive disorder.

**Supplementary Table 1. Characteristics of the GWAS summary data**

| **Phenotype** | | **Sample size** | **Data source** | **Units of effect size** | **Ancestry** | **Phenotype description** | **PubMed ID** |
| --- | --- | --- | --- | --- | --- | --- | --- |
| **Asthma** | |  |  |  |  |  |  |
|  | Childhood-onset (onset between 0-12 years) | 9433 cases and 318,237 controls | UKBB | OR | European | Self-reports of doctor-diagnosed asthma cases with onset at younger than 12 years | 31036433 |
|  | Age of onset | 37846 | UKBB | years | European | Age of onset of asthma | 31036433 |
| **Mental disorders** | |  |  |  |  |  |  |
|  | MDD | 45,591 cases and 97,674 controls | PGC | OR | European | Clinically diagnosed MDD without participants from UKBB and 23andMe cohorts | 29700475 |
|  | Bipolar disorder | 40,463 cases and 313,436 controls | PGC | OR | European | Clinically diagnosed bipolar disorder without participants from UKBB cohort | 34002096 |
|  | Schizophrenia | 53,386 cases and 77,258 controls | PGC | OR | European | Clinically diagnosed Schizophrenia | 35396580 |
|  | Anxiety/panic disorder | 2248 cases and 7992 controls | PGC | OR | European | Clinically diagnosed panic disorder | 31712720 |
|  | Autism | 18381 cases and 27969 controls | PGC | OR | European | Clinically diagnosed autism spectrum disorder | 30804558 |
|  | ADHD | 38,691 cases and 186,843 controls | PGC | OR | European | Clinically diagnosed ADHD | 36702997 |

UKBB: UK Biobank; PGC: Psychiatric Genomics Consortium; MDD: Major Depressive Disorder; ADHD: attention deficit hyperreactivity disorder; OR: odds ratio.

**Supplementary Table 2. The strengths of the instrumental variables used in the study**

| **Exposure** | **Outcome** | **nSNP** | **R2** | **F_stat** |
| --- | --- | --- | --- | --- |
| childhood onset asthma | ADHD | 16 | 0.004071695 | 83.72233819 |
| childhood onset asthma | Schizophrenia | 18 | 0.00437322 | 79.95465235 |
| childhood onset asthma | Anxiety | 18 | 0.004051594 | 74.05051311 |
| childhood onset asthma | Autism | 18 | 0.00437322 | 79.95465235 |
| childhood onset asthma | Bipolar disorder | 19 | 0.004478325 | 77.5749419 |
| childhood onset asthma | MDD | 18 | 0.004051594 | 74.05051311 |
| age of onset of asthma | ADHD | 12 | 0.022908467 | 73.91801784 |
| age of onset of asthma | Schizophrenia | 9 | 0.011069795 | 47.05834453 |
| age of onset of asthma | Anxiety | 12 | 0.020393828 | 65.63519502 |
| age of onset of asthma | Autism | 12 | 0.021960058 | 70.78910719 |
| age of onset of asthma | Bipolar disorder | 9 | 0.009803653 | 41.62260873 |
| age of onset of asthma | MDD | 12 | 0.020393828 | 65.63519502 |

MDD: Major Depressive Disorder; ADHD: attention deficit hyperreactivity disorder.

**Supplementary Table 3. Causal effects of childhood asthma on mental disorders with three MR approaches**

| **Exposure** | **Outcome** | **Method** | **OR (95% CI)** | **p** |
| --- | --- | --- | --- | --- |
| Childhood-onset asthma | MDD | Inverse variance weighted | 1.059 (1.025-1.095) | 5.72E-04 |
|  | MDD | Weighted median | 1.043 (0.996-1.093) | 7.64E-02 |
|  | MDD | MR Egger | 1.046 (0.974-1.123) | 2.34E-01 |
|  | Bipolar disorder | Inverse variance weighted | 1.065 (1.027-1.105) | 6.75E-04 |
|  | Bipolar disorder | Weighted median | 1.093 (1.042-1.145) | 2.41E-04 |
|  | Bipolar disorder | MR Egger | 1.117 (1.033-1.209) | 1.34E-02 |
|  | Schizophrenia | Inverse variance weighted | 1.026 (0.991-1.062) | 1.48E-01 |
|  | Schizophrenia | Weighted median | 1.044 (0.997-1.094) | 6.64E-02 |
|  | Schizophrenia | MR Egger | 1.055 (0.977-1.139) | 1.92E-01 |
|  | ADHD | Inverse variance weighted | 1.007 (0.974-1.042) | 6.78E-01 |
|  | ADHD | Weighted median | 0.993 (0.948-1.041) | 7.79E-01 |
|  | ADHD | MR Egger | 1 (0.928-1.077) | 9.97E-01 |
|  | Autism | Inverse variance weighted | 1 (0.944-1.059) | 9.97E-01 |
|  | Autism | Weighted median | 0.955 (0.894-1.019) | 1.63E-01 |
|  | Autism | MR Egger | 0.979 (0.859-1.115) | 7.51E-01 |
|  | Anxiety | Inverse variance weighted | 1.038 (0.875-1.232) | 6.65E-01 |
|  | Anxiety | Weighted median | 0.933 (0.78-1.117) | 4.53E-01 |
|  | Anxiety | MR Egger | 0.865 (0.6-1.247) | 4.48E-01 |
| Age of onset of asthma | MDD | Inverse variance weighted | 0.993 (0.988-0.998) | 3.27E-03 |
|  | MDD | Weighted median | 0.994 (0.987-1.001) | 8.66E-02 |
|  | MDD | MR Egger | 0.987 (0.976-0.999) | 6.51E-02 |
|  | Bipolar disorder | Inverse variance weighted | 0.999 (0.992-1.006) | 7.32E-01 |
|  | Bipolar disorder | Weighted median | 1 (0.991-1.009) | 9.72E-01 |
|  | Bipolar disorder | MR Egger | 0.987 (0.963-1.011) | 3.10E-01 |
|  | Schizophrenia | Inverse variance weighted | 0.996 (0.99-1.002) | 2.22E-01 |
|  | Schizophrenia | Weighted median | 0.997 (0.989-1.005) | 4.98E-01 |
|  | Schizophrenia | MR Egger | 1.004 (0.98-1.029) | 7.50E-01 |
|  | ADHD | Inverse variance weighted | 0.999 (0.995-1.004) | 7.73E-01 |
|  | ADHD | Weighted median | 1.001 (0.994-1.007) | 8.66E-01 |
|  | ADHD | MR Egger | 1.004 (0.991-1.017) | 6.02E-01 |
|  | Autism | Inverse variance weighted | 1.002 (0.994-1.011) | 5.54E-01 |
|  | Autism | Weighted median | 1.006 (0.997-1.016) | 2.03E-01 |
|  | Autism | MR Egger | 1.007 (0.984-1.031) | 5.57E-01 |
|  | Anxiety | Inverse variance weighted | 1.006 (0.987-1.025) | 5.27E-01 |
|  | Anxiety | Weighted median | 1.013 (0.986-1.04) | 3.49E-01 |
|  | Anxiety | MR Egger | 1.025 (0.976-1.076) | 3.51E-01 |

MDD: Major Depressive Disorder; ADHD: attention deficit hyperreactivity disorder.
